# Analysis of Corneal Surface Temperature Changes Following Fingertip, Knuckle, and Fingernail Eye Rubbing

**DOI:** 10.64898/2026.04.29.26351280

**Authors:** Metehan Karaatlı, Musacan Yalçın, Sertaç Eroğlu, Onur Özalp, Eray Atalay

## Abstract

**Purpose:** To characterize corneal surface temperature changes induced by different eye-rubbing techniques in healthy individuals and to investigate the factors influencing temperature change.

**Setting:** Eskisehir Osmangazi University

**Design:** Cross-sectional experimental study

**Methods:** This study included 93 healthy volunteers aged 19-29 years with no ocular pathology. Participants performed three eye-rubbing techniques-fingertip, knuckle, and fingernail rubbing-while corneal temperatures were recorded with a high-resolution thermal camera (FLIR A8200sc, Teledyne FLIR Systems Inc., Boston MA, USA). Subjects rubbed their eyes for 20 seconds with their dominant hand. Linear mixed-effects models were used to compare corneal temperature before and after eye rubbing and to examine the effect of covariates.

**Results:** All eye rubbing techniques significantly increased corneal temperature (fingertip: 1.02 +/-0.58 degrees Celsius, knuckle: 1.03 +/-0.54 degrees Celsius, fingernail: 1.12 +/-0.52 degrees Celsius; all p<0.001), with no significant differences between techniques (p>0.05). Age showed a negative correlation with corneal temperature increase across all rubbing methods (all unadjusted p<0.05), remaining significant only for the fingertip technique after FDR correction (p<0.001). IHA correlated positively with temperature increase for fingertip and knuckle rubbing after FDR adjustment (p= 0.003 and <0.001, respectively). The subgroup analysis indicated that approximately 0.6 degrees Celsius of every 1 degrees Celsius rise in corneal temperature could be attributed to eye closure alone, while the remainder was likely due to mechanical effects of eye rubbing.

**Conclusion:** Fingertip, knuckle, and fingernail rubbing each produced a transient but significant rise of approximately 1 degree Celsius in corneal temperature. Greater temperature elevation was associated with younger age and higher corneal asymmetry.

## INTRODUCTION

Eye rubbing is a common physiological response to conditions such as emotional stress, fatigue, exposure to dust and allergens, or ocular irritation associated with contact lens use.^1, 2^ Ocular surface diseases such as dry eye syndrome, allergic conjunctivitis, atopy, and meibomian gland dysfunction can trigger eye rubbing^3-5^. Eye rubbing can trigger or exacerbate biomechanical weakening of the cornea and is considered a causative factor in ectatic corneal disorders, most notably keratoconus.^5, 6^ In addition to direct mechanical insult, eye rubbing generates frictional heat that elevates corneal surface temperature and transiently reduces its biomechanical resistance. Repetitive habitual eye rubbing is therefore implicated in the onset and progression of ectatic corneal disease.^7-9^

There is no universal method of eye rubbing; individuals exhibit distinct patterns and techniques. A survey-based study identified four primary eye rubbing techniques—using the fingertips, knuckles, back of the hand, and base of the thumb—the first two of which accounted for over 90% of all responses among keratoconus patients, healthy individuals without contact lens wear, and healthy contact lens wearers.^10^ A subsequent study that recorded participants performing their habitual eye-rubbing motion reported that rubbing with the fingertip was most frequent (51%), followed by rubbing with the knuckle (44%) and the fingernail (6%). The same study quantified the mechanical forces applied to the eyelids using a high-precision balance and found marked variability across the rubbing techniques, with knuckle-type rubbing exerting significantly greater force (9.6 ± 6.3 N) compared to fingertip (4.3 ± 3.1 N) and fingernail (2.6 ± 3.3 N) techniques.^11^ These findings suggest that different eye rubbing styles impose variable mechanical stress on the cornea, and that certain techniques may be more detrimental to corneal biomechanical integrity and contribute more strongly to keratoconus development or progression

While the afore-mentioned study clearly shows that different eye rubbing techniques exert varying levels of mechanical forces, no study has yet quantified the resulting temperature changes on the corneal surface after these eye rubbing techniques. Therefore, this study aimed to quantitatively evaluate corneal temperature changes induced by the three most commonly described eye-rubbing techniques in the literature, using high-resolution thermal imaging, and to identify potential factors associated with temperature variation.

## METHODS

This cross-sectional study was conducted at the cornea clinics of Eskişehir Osmangazi University Hospital between September 2023 and November 2023 on healthy volunteers aged 18 to 30 years with no ocular pathologies. The study was approved by the Ethical Board of Non-Interventional Clinical Research of Eskişehir Osmangazi University (Decision no: 18; date: 25.07.2023) and was conducted in adherence to the tenets of the Declaration of Helsinki. Written informed consent was obtained from all participants prior to inclusion in the study.

### Participants

Subjects were prospectively recruited and underwent a comprehensive ophthalmologic examination, including corneal tomography (Pentacam, Oculus Optikgeräte GmbH, Wetzlar Germany), optical biometry (Lenstar LS 900, Haag-Streit AG, Koeniz, Switzerland.), and autorefraction (ARK-1, Nidek Co., Ltd., Gamagori, Japan), to confirm eligibility. Exclusion criteria comprised known chronic ocular disease (including keratoconus), history of ocular surgery or trauma, signs or symptoms of allergic conjunctivitis or ocular inflammation, evidence of dry eye, myopia > –6.00 D or axial length >26 mm, contact lens wear on the study day, and use of any topical ophthalmic medication within two weeks prior to participation.

### Experimental Procedure

All experiments were conducted under standard room conditions without strict environmental control, and ambient temperature was recorded to evaluate its potential influence on post-rubbing corneal temperature changes. Each participant underwent a 5-minute acclimatization period before measurement to allow stabilization of ocular surface temperature. Participants then positioned their heads on a combined head- and chinrest at a fixed 50 cm distance from the thermal camera, corresponding to the manufacturer-specified focal length.

Baseline corneal temperature was recorded over a 20-second resting period, during which the mean value was calculated. Participants subsequently rubbed their eyes for 20 seconds using one of three predefined techniques—fingertip, knuckle, or fingernail rubbing **(Figure 1)** using their dominant hand. Immediately after rubbing ceased, corneal temperature was measured from the first thermal frame in which the corneal surface was visible following full eye reopening. Each rubbing technique was applied in a randomized order, with a 10-minute rest interval between sessions to allow temperature stabilization and prevent carry-over effects.

**Figure 1.**
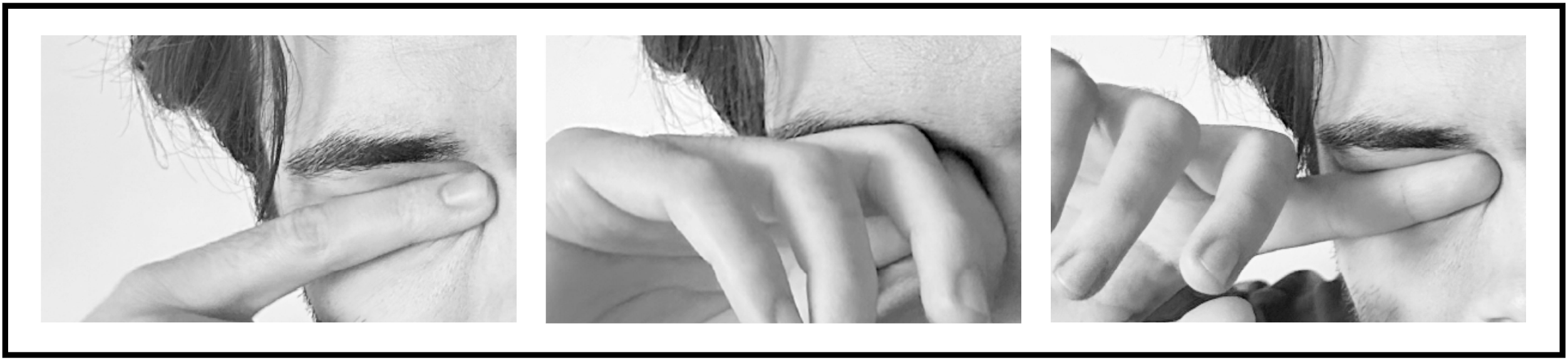
Demonstration of the three eye rubbing techniques evaluated in the study: (left panel) fingertip, (middle panel) knuckle, and (right panel) fingernail rubbing.

Recordings at 20 frames per second were made using a high-resolution thermal camera (FLIR A8200sc, Teledyne FLIR Systems Inc., Boston MA, USA). The camera has a native resolution of 1024 × 1024 pixels, a pixel pitch of 18 µm, and a thermal sensitivity <0.02 °C. Temperature was measured in a 3-mm region of interest centred on the corneal apex while the patient was fixating on a fixed focal point.

### Sample Size Calculation and Statistical Analysis

The expected mean temperature increases were 0.5 °C for fingernail rubbing, 1.0 °C for fingertip rubbing, and 1.5 °C for knuckle rubbing relative to baseline, assuming a common standard deviation of 1.0 °C. These expectations were derived from the progressively increasing rubbing forces reported by Hafezi et al: 2.6 ± 3.3 N for fingernail, 4.3 ± 3.1 N for fingertip, and 9.6 ± 6.3 N for knuckle rubbing, approximately doubling with each method.^11^ Based on these assumptions, an a priori power analysis using G*Power v3.1 (one-way ANOVA, fixed effects) with *f* = 0.41, α = 0.05, and power = 0.95 indicated that a total of n = 96 participants was required.

All analyses were performed using R version 4.4.3. Linear mixed-effects models were used to compare corneal temperature before and after eye rubbing and to examine the effect of ambient temperature. We also evaluated the influence of age, sex, refraction and ocular and corneal parameters both from biometry and Pentacam HR, including axial length, spherical equivalent, keratometry, pachymetry, corneal volume, irregularity indices, Kmax, BAD-D, best-fit sphere, progression indices, and Ambrosio relational thickness, for each rubbing technique using both linear mixed-effects models and Pearson correlation analyses. To distinguish potential thermal transfer resulting from increased proximity between the palpebral conjunctival vasculature and the cornea during eye rubbing, 30 individuals who had also closed their fellow eye were identified during post hoc thermal video analysis. The difference in corneal surface temperature between the rubbed and fellow eyes was then compared in this subgroup using paired samples t test. A *p* value < 0.05 was considered statistically significant, and false discovery rate (FDR) correction was applied for multiple comparisons.

## RESULTS

This cross-sectional study included 93 healthy participants (mean age: 22.8 ± 1.6 years), of whom 49 (52.7%) were male and 44 (47.3%) were female. Descriptive statistics for the axial length, spherical equivalent and corneal parameters are presented in **Table 1**. Baseline corneal temperatures were comparable across rubbing conditions (Fingertip: 34.3 ± 0.9 °C; Knuckle: 34.3 ± 0.8 °C; Fingernail: 34.3 ± 0.8 °C; overall mean = 34.3 ± 0.8 °C). Following fingertip, knuckle, and fingernail rubbing, corneal temperatures increased by 1.02 ± 0.58 °C, 1.03 ± 0.54 °C, and 1.12 ± 0.52 °C, respectively (all p < 0.001). Ambient temperature did not significantly affect corneal temperature change for any of the eye rubbing techniques (all p <0.05), indicating that room temperature variations had minimal influence on the results.

**Table 1.**
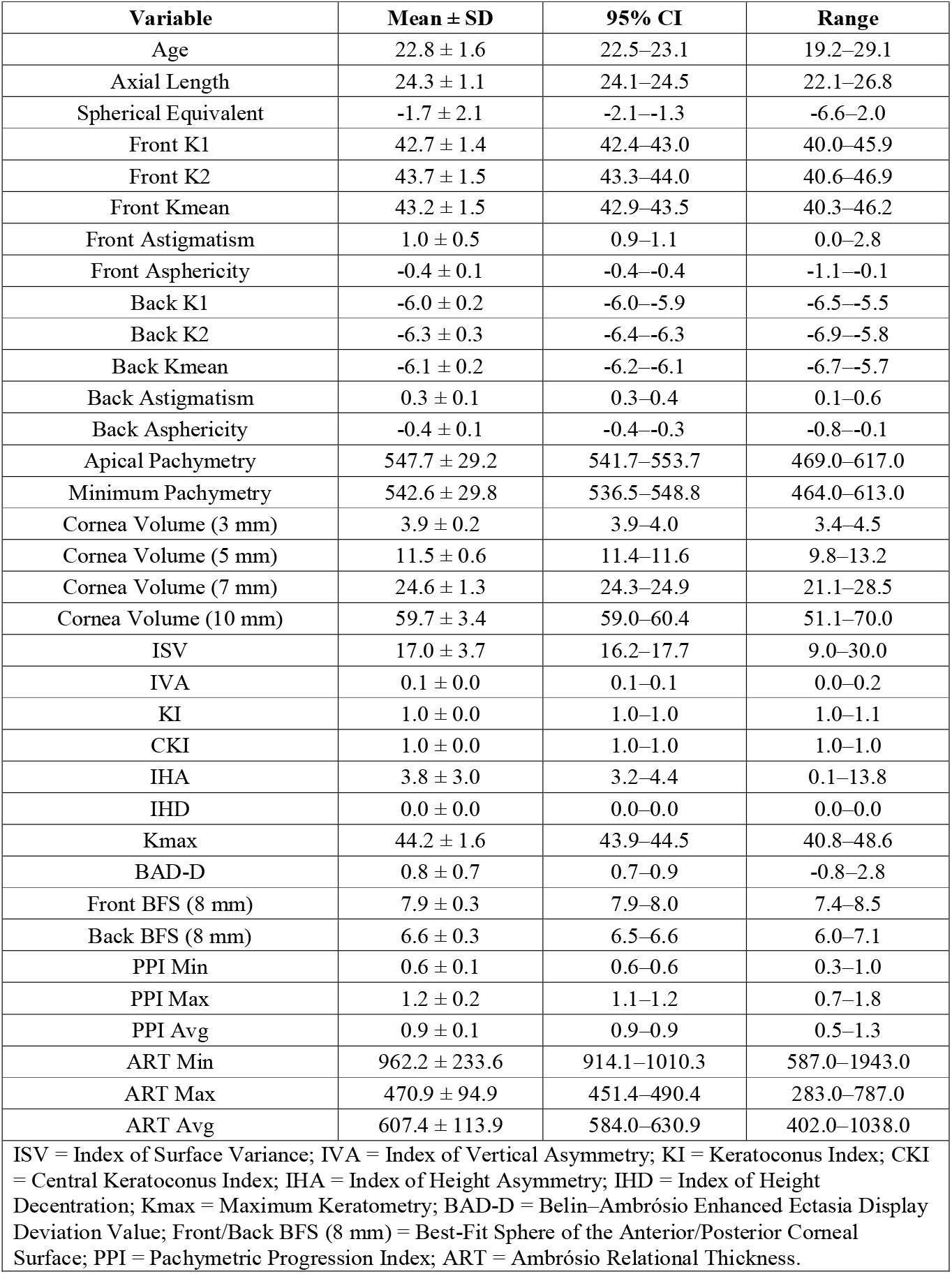
Descriptive statistics for the axial length, spherical equivalent and corneal parameters.

**Figure 2** shows the violin plot illustrating the distribution of corneal temperature change (Δ Temperature) across the three rubbing techniques. All rubbing techniques produced a positive change in corneal temperature. Notably, in the fingernail rubbing condition, no subjects exhibited a decrease in temperature, whereas a few cases in the knuckle and fingernail rubbing conditions showed minimal or no increase. Nonetheless, there were no statistically significant differences between techniques (all p > 0.05). In the subgroup analysis of 30 participants who incidentally closed their fellow eye during rubbing, the rubbed eye demonstrated a significantly greater corneal temperature increase (1.20 ± 0.56 °C) than the fellow eye (0.72 ± 0.40 °C; p < 0.001, **Figure 3**).

According to the linear mixed-effects model, age was negatively associated with corneal temperature increase in all models (all unadjusted *p* < 0.05, **Table 2**); however, when *p* values were adjusted for multiple comparisons, this association remained statistically significant only in the overall and fingertip rubbing models (*p =* 0.002 and *p* < 0.001, **Table 2**). Furthermore, the index of height asymmetry (IHA) consistently demonstrated a significant but a weakly positive association with corneal temperature increase across all conditions (all unadjusted *p* < 0.05, **Table 2**). After adjustment for multiple testing, this association remained statistically significant in the combined, fingertip, and knuckle rubbing models (adjusted *p* < 0.05, **Table 2**), whereas in the fingernail rubbing model, significance was lost (adjusted *p* = 0.08, **Table 2**). While no other parameters showed significant associations in the remaining models, in the fingertip rubbing model, the index of surface variance (ISV) and ART-Min remained significant predictors in the fingernail rubbing model (adjusted *p* = 0.04 and 0.02, respectively, **Table 2**).

**Table 2.**
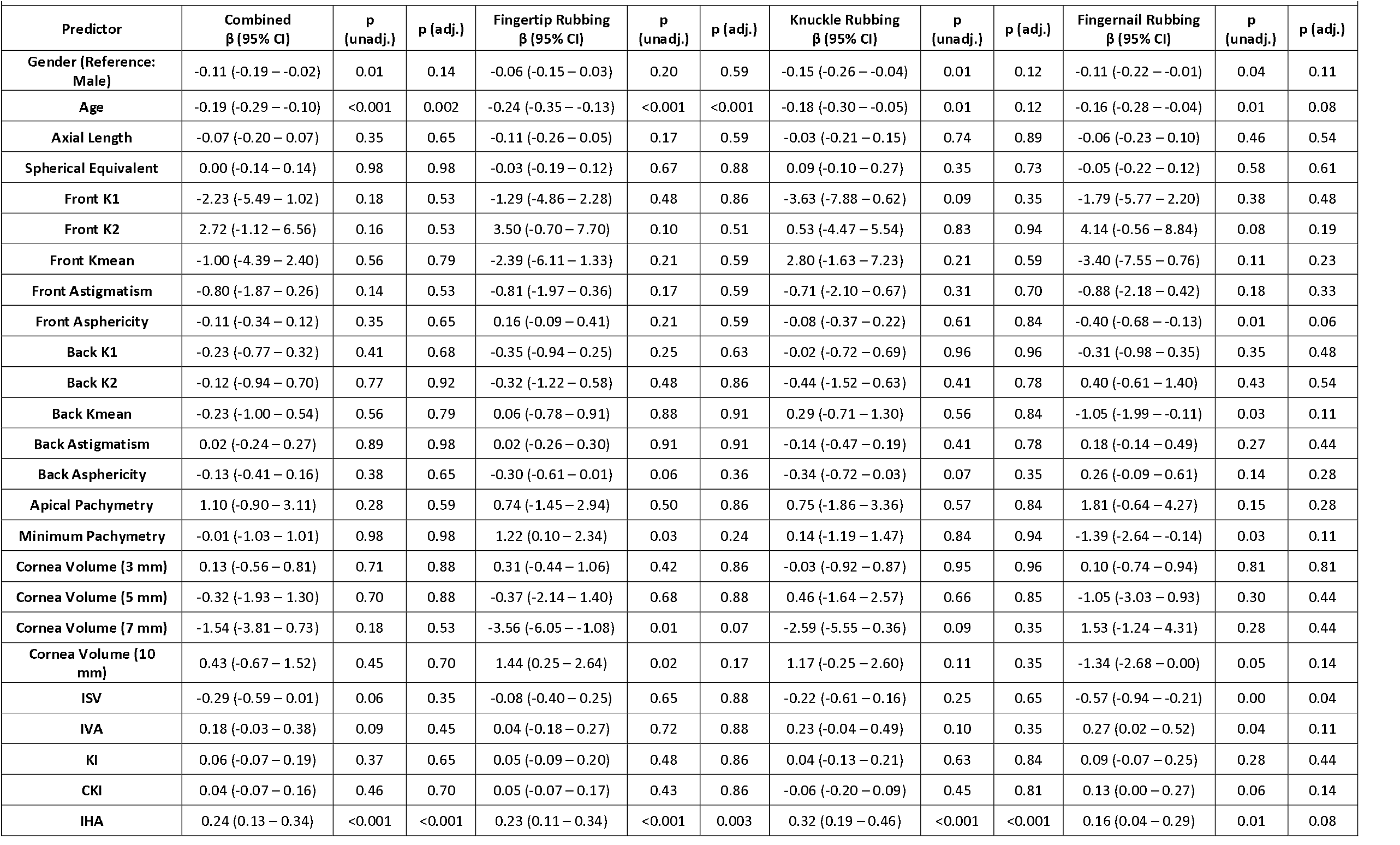
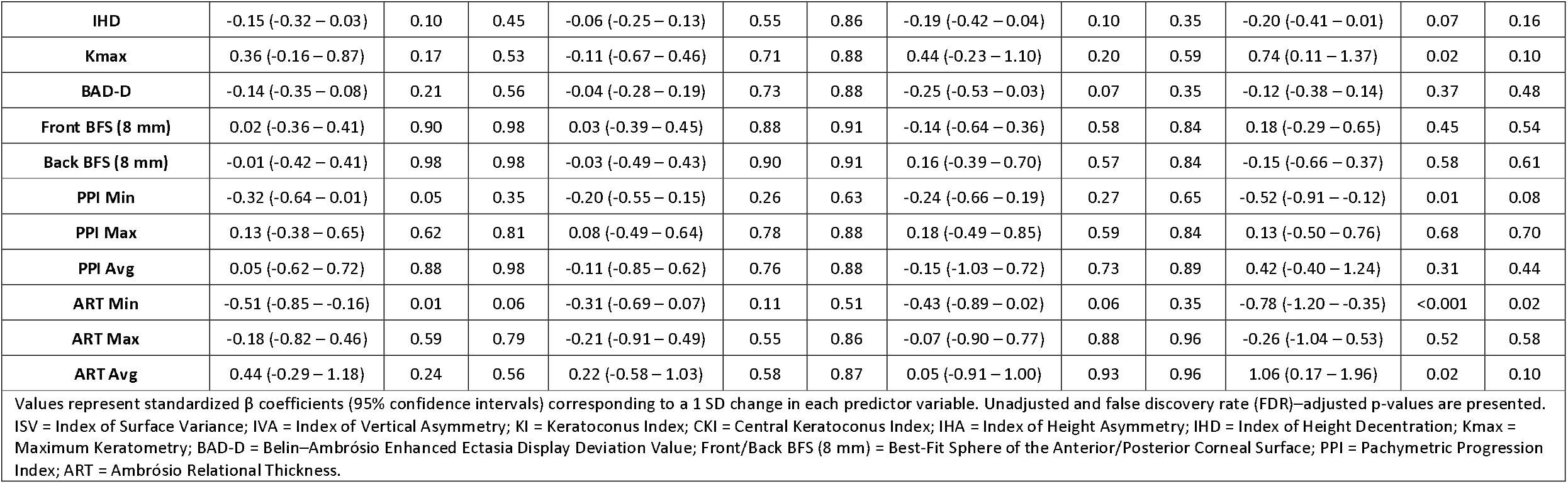
Linear mixed-effects model results showing predictors of corneal temperature increase after eye rubbing.

**Figure 2.**
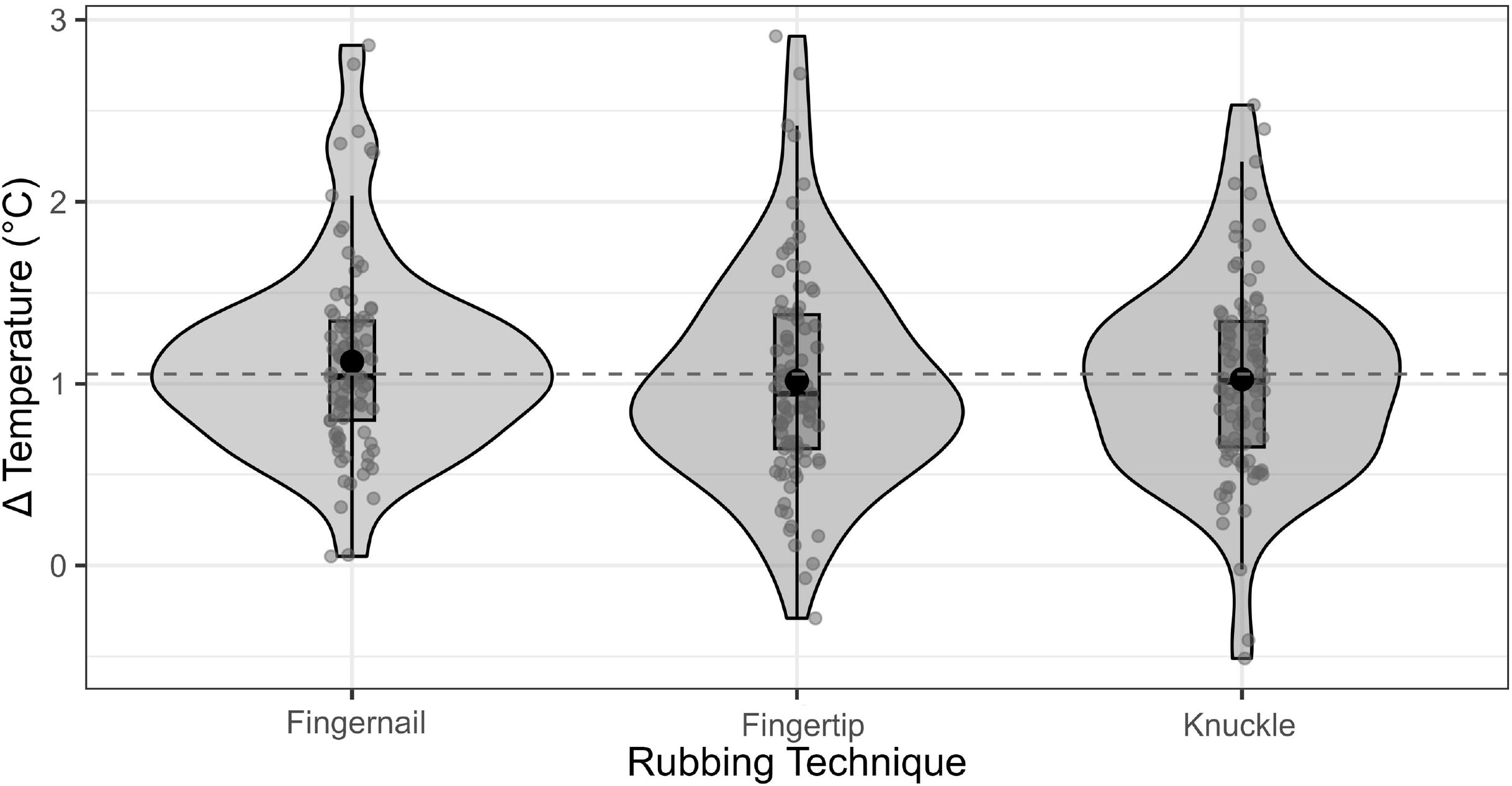
Violin plots showing the distribution of corneal temperature change (Δ Temperature, °C) after fingertip, knuckle, and fingernail rubbing. Each plot displays individual values (gray dots), the median and interquartile range (box), and the group mean (black dot). The dashed line indicates the overall mean temperature change.

**Figure 3.**
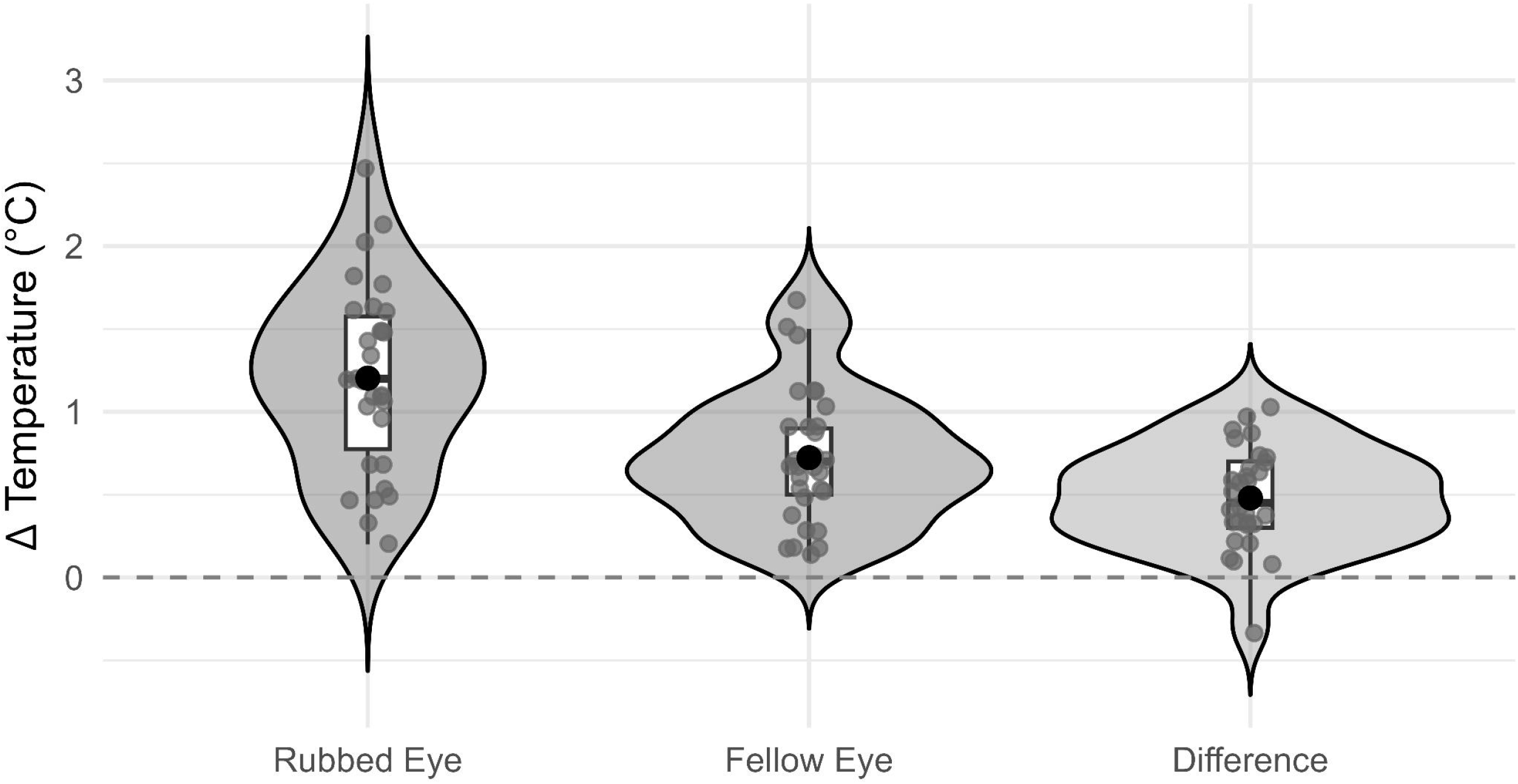
Comparison of corneal temperature changes between rubbed and fellow eyes. Violin plots illustrate the distribution of corneal temperature increases measured after eye rubbing in the rubbed eye and the fellow (non-rubbed) eye, as well as the inter-eye difference (rubbed – fellow). Each plot displays individual values (gray dots), the median and interquartile range (box), and the group mean (black dot).

The correlations between corneal temperature changes induced by three different eye rubbing techniques and spherical equivalent, axial length, and Pentacam-derived parameters were evaluated **(Supplementary Figure)**. Correlation analyses revealed a statistically significant positive association between corneal temperature elevation and IHA across all three eye-rubbing modalities (all unadjusted *p* < 0.05), which only remained significant after FDR correction in the combined and knuckle rubbing models. Weak but consistent positive correlations were also noted between corneal temperature increase and corneal volume parameters (3–10 mm diameters) and front asphericity in the fingertip model, however these correlations did not hold true after FDR correction.

## DISCUSSION

In this cross-sectional study, we used high-resolution thermal imaging to measure changes in the temperature of the corneal surface induced by three commonly used eye rubbing techniques in healthy participants. All eye rubbing techniques examined (fingertip, knuckle and fingernail) resulted in a statistically significant increase in corneal temperature (fingertip 1.02 ± 0.58 °C, knuckle 1.03 ± 0.54 °C, fingernail 1.12 ± 0.52 °C; all *p* < 0.001), with no significant difference between techniques (*p* > 0.05). According to the linear mixed-effects model, age was negatively associated with corneal temperature increase in all models (all unadjusted *p* < 0.05, **Table 2**); after FDR correction this remained significant only for the overall and fingertip models (adjusted *p =* 0.002 and *p* < 0.001, **Table 2**). Furthermore, IHA was positively correlated with corneal temperature increase (unadjusted *p* < 0.05, **Table 2**); after FDR correction this remained significant for the overall, fingertip, and knuckle models (adjusted *p* < 0.05, **Table 2**), while the fingernail model lost significance though a positive trend persisted (adjusted *p* = 0.079, **Table 2**). Heat map analysis supported these findings, showing a consistent positive correlation between IHA and corneal temperature increase across the eye rubbing techniques evaluated **(Supplementary Figure)**.

An increase in corneal surface temperature induced by eye-rubbing has multiple biomechanical and biochemical ramifications for the cornea, all of which contribute to the risk of ectasia development. First, elevated corneal temperature coupled with the shear forces generated during eye rubbing transiently alters viscoelastic properties of the cornea, leading to a reduction in its bending resistance.^2, 12^ In line with this, studies have shown progressive reduction in viscoelastic properties of the cornea (namely corneal resistance factor [CRF] and corneal hysteresis [CH]) after 20 seconds of self-inflicted eye rubbing.^13^ Second, temperature elevation enhances the activity of degradative enzymes.^7^ Given that eye rubbing concurrently induces local release of cytokines and inflammatory mediators, the resulting biochemical alteration further aggravates stromal degradation contributing to corneal biomechanical weakening.^1^ Third, the mechanical impact of eye rubbing directly imposes shear stress on the cornea while transiently elevating intraocular pressure, reportedly as high as high as 135 mmHg, which further compromises the mechanical integrity of the cornea.^14^

Among the factors contributing to the rise in corneal temperature during eye rubbing, the primary mechanism may be the increased proximity of the highly vascular palpebral conjunctiva to the corneal surface.^7, 15, 16^ This is consistent with reported measurements showing that, while normal corneal temperature averages approximately 34–35 °C at room temperature, the upper eyelid conjunctiva reaches much higher values, around 36.2 ± 0.6 °C.^7, 17, 18^ Hence, the increased contact of a warmer palpebral conjunctiva with a cooler corneal surface could be a major driver of transient increase in corneal temperature with eye rubbing.^7^ Additionally, the closure of eyelids during rubbing may reduce heat loss through tear evaporation, convection, and radiation, and promoting transient heat accumulation.^7, 19^ Finally, part of the temperature elevation may result from frictional heat generation, viscous damping, and heat dissipation associated with the viscoelastic properties of the cornea.^20^ To isolate the effect of passive thermal transfer from the palpebral conjunctiva and inhibition of heat loss, we performed a subgroup analysis of individuals who incidentally closed their fellow eye during rubbing **(Figure 3)**. This analysis indicated that approximately 0.6 °C of every 1 °C increase in corneal temperature could be attributed to conjunctival contact and heat retention alone, whereas the additional rise observed in the rubbed eye likely resulted from frictional heating.

Our study revealed a negative correlation between age and the increase in corneal temperature induced by eye rubbing. In younger individuals, the cornea exhibits relatively more hydrated collagen with lower degree of cross-linking hence greater viscoelasticity, whereas in older individuals, the cornea become stiffer and less flexible.^21, 22^ Because younger corneas exhibit greater viscoelasticity, their viscous damping capacity is higher, allowing more energy to be dissipated as heat from eye rubbing. Another possible explanation for the negative correlation between age and corneal-temperature rise after eye-rubbing involves age-related reductions in ocular tissue temperatures. Previous work shows that the mean temperatures of the cornea, limbus, sclera and outer canthus decline with increasing age.^23, 24^ While no study has yet evaluated, it is conceivable that the palpebral conjunctiva also undergoes an age-related decline in baseline temperature, resulting in reduced heat transfer with eye rubbing in older individuals. Collectively, these findings suggest that younger eyes may be more susceptible to thermal effects induced by eye rubbing.

Among the ocular parameters, a significant positive correlation was found between the corneal temperature increase induced by eye rubbing techniques and IHA. IHA, a Pentacam-derived parameter, represents the average difference in elevation values between the superior and inferior regions of the cornea, using the horizontal meridian as the axis of symmetry and is elevated in ectatic corneal diseases.^25, 26^ Elevated IHA values indicate that the cornea exhibits anterior corneal surface asymmetry. During eye rubbing, this irregular surface may lead to nonuniform contact between the palpebral conjunctiva and corneal surface, potentially increasing friction and, consequently, heat generation.

This study has several limitations. First, it was conducted on healthy volunteers, and the magnitude of corneal temperature increase after eye rubbing may differ in individuals with corneal pathology. Second, the ambient temperature was not strictly controlled during the experiments. Nevertheless, we attempted to account for this variable by recording ambient temperature for each participant and assessing its effect through statistical analysis. Although previous studies report that ocular surface temperature changes only minimally with room temperature (0.15 to 0.2°C per degree increase in room temperature), we did not observe a significant influence of ambient temperature on the temperature change induced by eye rubbing.^16, 27^ Third, as expected, the amount of force applied to the eyelid during rubbing may not be comparable across the subjects for each rubbing method. However, we did not observe a difference amongst the three eye rubbing methods, each associated with varying degrees of applied force, suggesting that applied force may not be a major determinant of the observed temperature rise.^11^ Furthermore, our measurement of corneal temperature was only limited to the central 3 mm region of interest; therefore it is unknown whether other corneal regions exhibit a different temperature profile after eye rubbing. Lastly, we did not measure finger temperature, which has been shown to correlate with corneal temperature.^28^ However, within the range of normal body temperature under standard room conditions, we do not anticipate that variations in finger temperature would have significantly influenced the temperature change induced by eye rubbing.

In conclusion, this study quantitatively assessed the impact of common eye rubbing techniques on corneal temperature, demonstrating that all techniques induced a significant temperature increase of approximately 1°C. Repetitive eye rubbing may compromise corneal biomechanics through both mechanical trauma and thermal effects, potentially contributing to keratoconus pathogenesis. Future research should further investigate the molecular mechanisms and clinical significance of this relationship.

## ACKNOWLEDGEMENTS

The authors sincerely thank medical students Zeynep Serra Özler, Taylan Can Özer, and Mert Egemen Çalışkan for their valuable assistance in performing the experimental procedures and data collection.

## Data availability

The dataset created and analysed during the current study is available from the corresponding author on reasonable request.

## VALUE STATEMENT

### What Was Known

- Different eye-rubbing techniques apply markedly different mechanical forces, with knuckle-type rubbing exerting the greatest load on the cornea.
- Friction during eye rubbing generates heat that elevates corneal surface temperature and transiently reduces corneal biomechanical resistance.

### What This Paper Adds

- Twenty-second eye rubbing caused an average corneal temperature increase of about 1 °C, with roughly 60% of the rise attributable to eye closure and the remainder to friction.
- Age showed an inverse correlation with corneal temperature elevation.
- Higher temperature increases were observed in more asymmetric corneas.

## FIGURE LEGENDS

**Supplementary Figure:** Correlation heatmap showing Pearson correlation coefficients (r) between corneal temperature change (Δ Temperature) and the parameters assessed for each rubbing technique and the combined dataset. Red indicates positive correlations, while blue indicates negative correlations. Asterisks (*) denote statistically significant correlations (p < 0.05), and daggers (†) indicate correlations that remained significant after false discovery rate (FDR) correction.

