## Supplementary figures and images for "Analysis of Corneal Surface Temperature Changes Following Fingertip, Knuckle, and Fingernail Eye Rubbing"

### Supplementary Figure

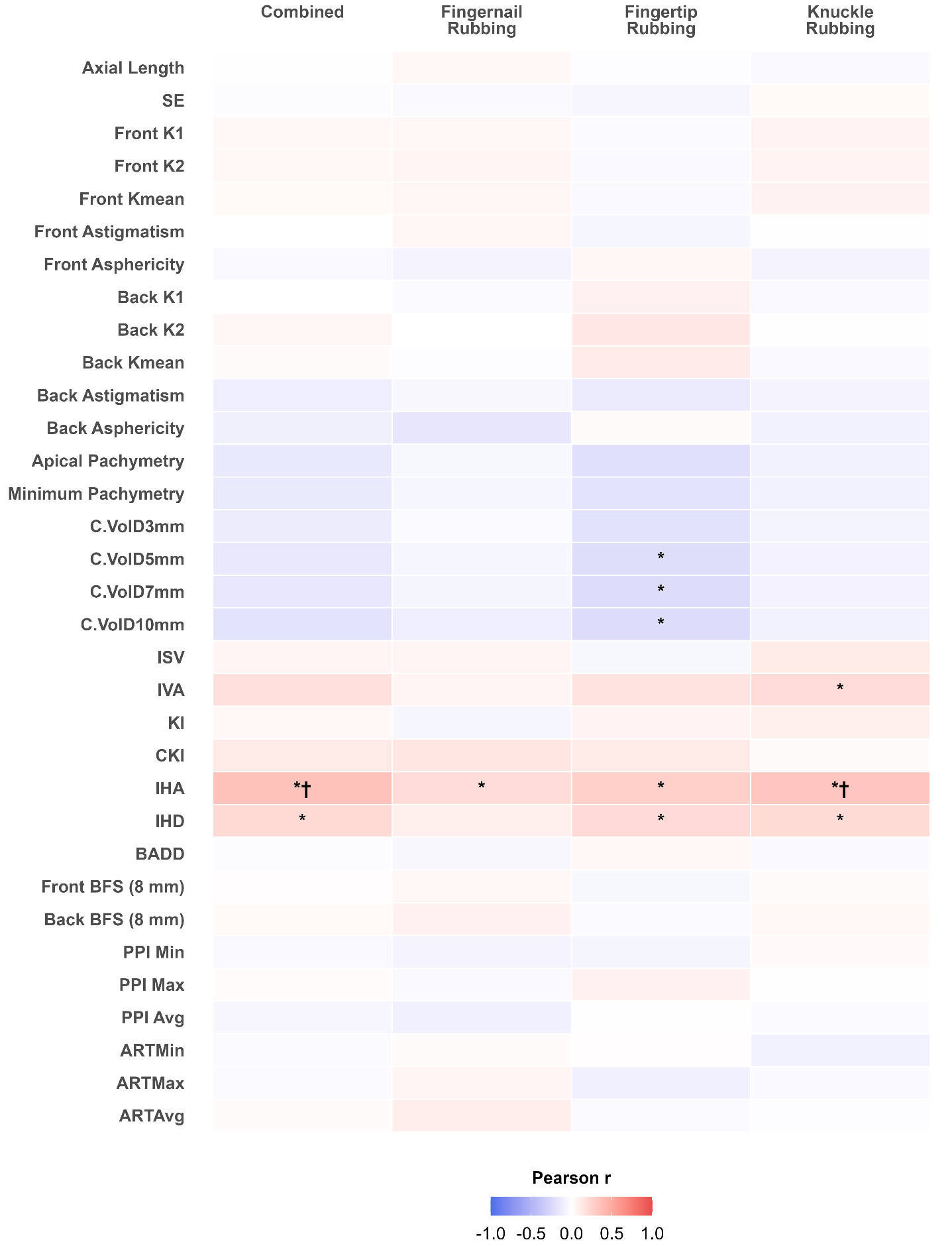
